# A cross-sectional study exploring the justification of opportunistic breast cancer screening in Thailand

**DOI:** 10.1101/2021.11.29.21266992

**Authors:** Bhoowit Lerttiendamrong, Lertpong Satapongpeera, Mawin Vongsaisuwon

## Abstract

**Objective:** Breast cancer is currently the most common malignant disease in Thailand. The present study aims to evaluate the most beneficial method of breast cancer screening in different breast densities by analyzing the benefits of screening mammography with additional breast ultrasonography classified by breast density.

**Method:** 49 middle-aged and elderly Bangkokian women who had undergone both mammography and ultrasonography were picked at random for analysis. BI-RADS scores were assigned based on mammography results alone and based on combined mammography and ultrasonography results. Concordance/discordance rates between the 2 radiographic techniques were compared in women stratified based on their breast densities.

**Results:** All of our participants were given a score between BIRADS 1 and 3, while over 40% of participants are in the BIRADS 2 category. 60% of subjects with extremely dense breasts benefit from screening mammography with additional breast ultrasonography, while only 50% of samples with heterogeneous density and 34.21% samples with heterogeneous fibroglandular breasts benefit from the extra intervention.

**Conclusion:** Our study concludes that women with higher breast density are more likely to benefit from screening using ultrasonography in addition to mammography as opposed to mammography screening alone. We recommend both mammography and ultrasonography for initial breast cancer screening. For follow-up visits, we suggest the screening method in accordance with breast density, using ultrasonography alone for women with high breast densities and mammography for women with heterogeneously dense breasts.

## Introduction

Breast cancer is the most common malignant disease among women in both developing and developed countries^1^. As of 2018, breast cancer accounts for 24% of new cancer cases and 15% of cancer deaths globally^2^. According to the 2011-2015 cancer registry conducted in Bangkok, Thailand, breast cancer is also the most prevalent cancer, comprising 29.4% of all cancer cases^3^. According to Virani et al., age-standardized incidence rate of breast cancer in 2012 in Thailand stands at 26.2 per 100,000 people^4^. Breast cancer is linked with westernized lifestyle and economic development^3^. As such, breast cancer incidence is projected to increase both globally and in the Asia-Pacific region. Global breast cancer incidence is increasing at a steady rate of 0.5% whilst incidence within the Asia-Pacific region is reporting an annual increase of 3-4%^5^.

Statistics have shown that between 1985 and 2015, survival in metastatic breast cancer has significantly improved with HER2 positive subset showing the most notable differences. The HER2 positive subtype improved survival resulted from the availability of transtuzumab and new cytotoxic agents^6^. Although breast cancer treatment is improving dramatically within the last few decades, breast cancer screening is lagging behind. Breast cancer screening, namely mammography, has been recommended for decades^7^, and still is the main breast cancer screening modality as of 2019^8^. Breast Imaging-Reporting and Data System score (BI-RADS score), established in 1995 and completed with ultrasonography interpretation in 2003, was set up as a quality assurance to homogenize the collected data by mammography and ultrasonography reports. BI-RADS score is still in use in most countries as of today^9^.

Current recommendations from the National Comprehensive Cancer Network (NCCN), published in 2018, differentiates breast cancer screening modality into 3 groups: basic, core and enhanced resources. Basic resources, aiming to improve-specific outcomes, recommend clinical encounters without the need for breast imaging. Core resources, providing major improvements in disease outcome without financial prohibition, recommend the use of ultrasonography alone. As for enhanced resources, the NCCN recommends diagnostic mammography with the use of screening mammography in high-risk cases^10^. The following recommendation by NCCN is in accordance with the American Cancer Society 2015 guideline for breast cancer, which recommends regular screening mammography at age 45 years in an average risk woman. Women between the age of 45 and 54 should be screened annually, whilst women 55 years and older should continue screening annually or transition to biennial screening^11^. However in women of Asian descent, a concern has been raised about accuracy of breast cancer detection as a result of high breast density as opposed to Western women^4^. In the status quo, guidelines and research publications within East Asian countries have suggested contradictory action. A retrospective study in women with breast cancer rural China concluded that detection of breast cancer using ultrasonography is more sensitive than mammography in women with high-density breasts, thus recommending ultrasonography for breast cancer screening^12^. On the other hand, the Japanese Breast Cancer Society Clinical Practice Guidelines of 2018 advises against using ultrasonography as an adjunct to breast cancer screening in the general population as there are currently no studies that have shown that the following intervention will reduce the breast cancer mortality rate^13^. Moreover, Okonkwo et al. showed that the cost-effectiveness of clinical breast examination in India compares favorably with the use of mammography in Western countries^14^.

Evidence suggests that Northeast Asian nations have incorporated recommendations from multiple randomized controlled trials into their clinical guideline. However, this is not the case with Thailand and other Southeast Asian nations. Research within Thailand reveals that the cost-effectiveness for once-in-a-lifetime breast cancer screening using mammography accounts for over 1.8 million Thai baht (approximately 57,000 US dollar using an exchange rate of 31.281 Baht per US dollar, exchange rate taken from Bank of Thailand on 1^st^ April 2021) per 1 year of quality-adjusted life year (QALY) in women between the ages of 40 and 49^15^. Thailand’s gross domestic product (GDP) per capita in fiscal year 2020 was 7,806.7 US dollars, referencing the data from the World Bank. As reflected from the following data, the cost of population-based breast cancer screening using mammography in Thailand is unjustifiable comparatively to the QALY gained by such screening examination, such mass screening, will adds a significant burden to the government financial status. To the best of our knowledge, there is currently no research conducted in Thailand or other Southeast Asian nations that investigates the difference between breast cancer detected by mammography alone versus mammography with ultrasonography. We identified the discordance between current breast cancer screening practice in Thailand and the lack of evidence supporting such practice. On 24^th^ March 2021, we performed a routine breast cancer screening event in Thai women using both mammography and ultrasonography. Within the span of 2 weeks since the opportunistic breast cancer screening event, we aim to explore the incidence of breast cancer characterized by Breast Imaging-Reporting and Data System score (BI-RADS score) sorted by age group and breast composition in middle-aged Thai women using adjunct ultrasonography with mammography and concordance rates between the two modalities. We hypothesize that additional ultrasonography might provide better detection rates for breasts with higher BI-RADS score.

## Methods

This research is conducted in Bangkok, capital city of Thailand, comprising over 10 million residents with the majority of inhabitants young or middle-aged adults. We randomly selected 49 middle-aged and older females currently living in Bangkok who are considered healthy without chronic illnesses. Middle-aged females in our study refer to females 36-55 years old whilst older females are those who are 56 years old and older. As for our 49 participants, their age range varies from 33 to 60 years old. Every participant had undergone both mammography and ultrasonography on 24^th^ March 2021. Their imaging results are assigned BI-RADS scores for each of the following: mammographic findings and ultrasonography findings. Error has been minimized by using the same mammography and ultrasonography device for all subjects. Furthermore, both mammographic findings and ultrasonographic findings are also interpreted by a single radiologist for all participants.

Subjects are then classified into 3 groups based on their breast density: heterogeneous fibroglandular density, heterogeneous dense and extremely dense breast. Results for each of the 3 groups are then interpreted by observing the concordance of BI-RADS score detected by mammography alone versus mammography with breast ultrasonography.

## Results

The participants’ ages ranged from 33 to 60 years with a mean age of 48.76 years. Participants are sorted by age into 6 age groups with an age interval of 5 years starting from 31-35 to 56-60. The age bracket with the greatest number of participants was the 56-60 bracket, comprised of 12 participants, while the age group with the least subjects was the 31-35 bracket with 3 subjects.

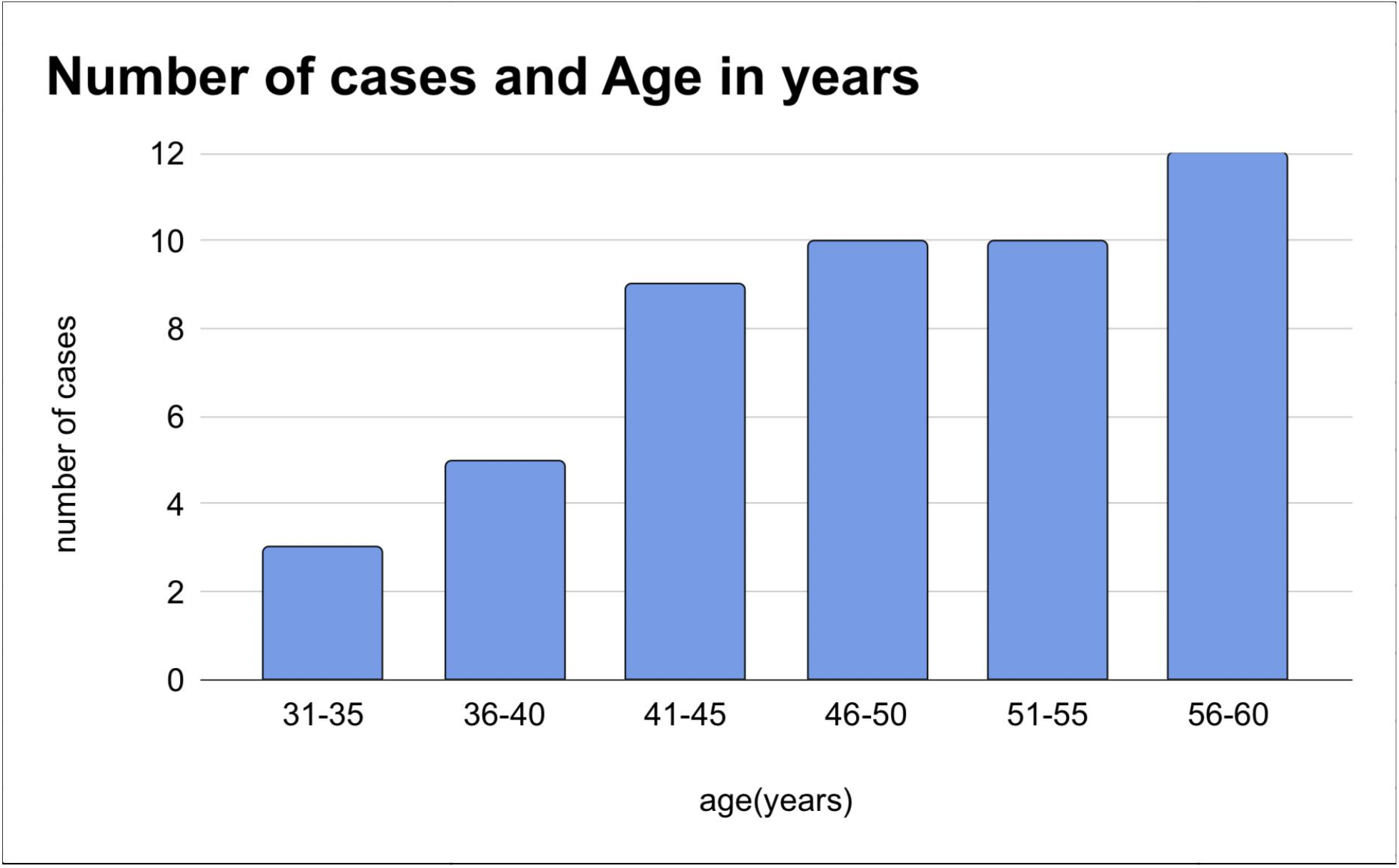

From the total of 49 participants, BI-RADS score using both mammographic and ultrasonographic findings are as follows: BI-RADS 1 with 19 subjects (38.78%), BI-RADS 2 with 20 subjects (40.82%), BI-RADS 3 with 10 subjects (20.41%). None of the participants had a BI-RADS 4 or 5 lesion.

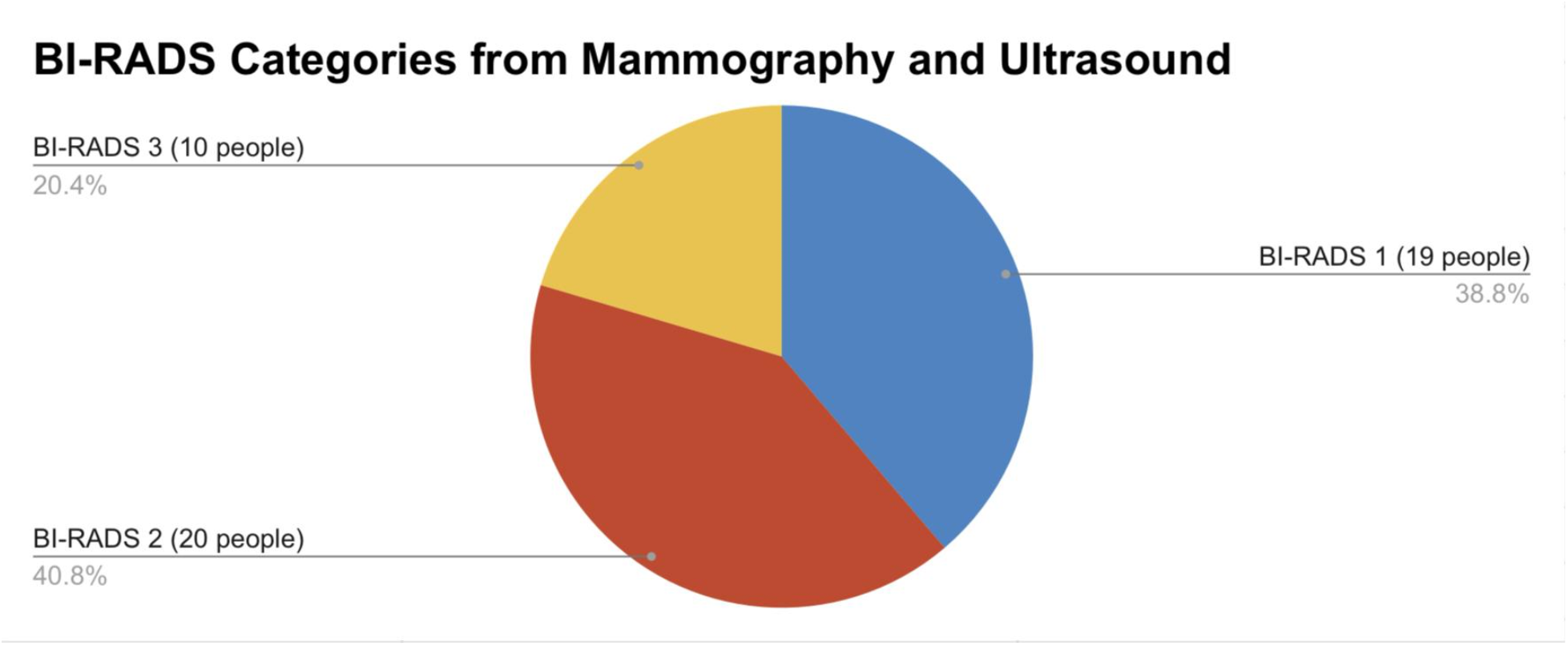

Participants are classified by their breast density into 3 groups: heterogeneous fibroglandular density, heterogeneous dense, and extremely dense breast. Most participants fall into the heterogeneous fibroglandular density category with 38 people, accounting for 77.55%.

Heterogeneous density and extremely dense breasts account for 6 (12.24%) and 5 (10.20%), respectively.

Each group of participants stratified by their breast density was then categorized by BI-RADS score concordance/discordance of mammogram alone versus mammogram plus ultrasound. Out of 38 participants with heterogeneous fibroglandular breast density, 29 participants (65.79%) had results in concordance between BI-RADS score assigned from mammogram alone and the score assigned using mammogram with adjunct breast ultrasound. On the other hand, 9 participants (34.21%) show a higher BI-RADS score if ultrasonography is added on top of mammogram. As for 6 participants with heterogeneous dense breasts, 3 participants (50%) show concordance while the other 3 participants show discordance between the BI-RADS score interpreted by mammogram versus that interpreted by combined mammogram and ultrasound. In the extremely dense breast group, only 2 participants out of 5 or 40% show concordance between the 2 imaging techniques.

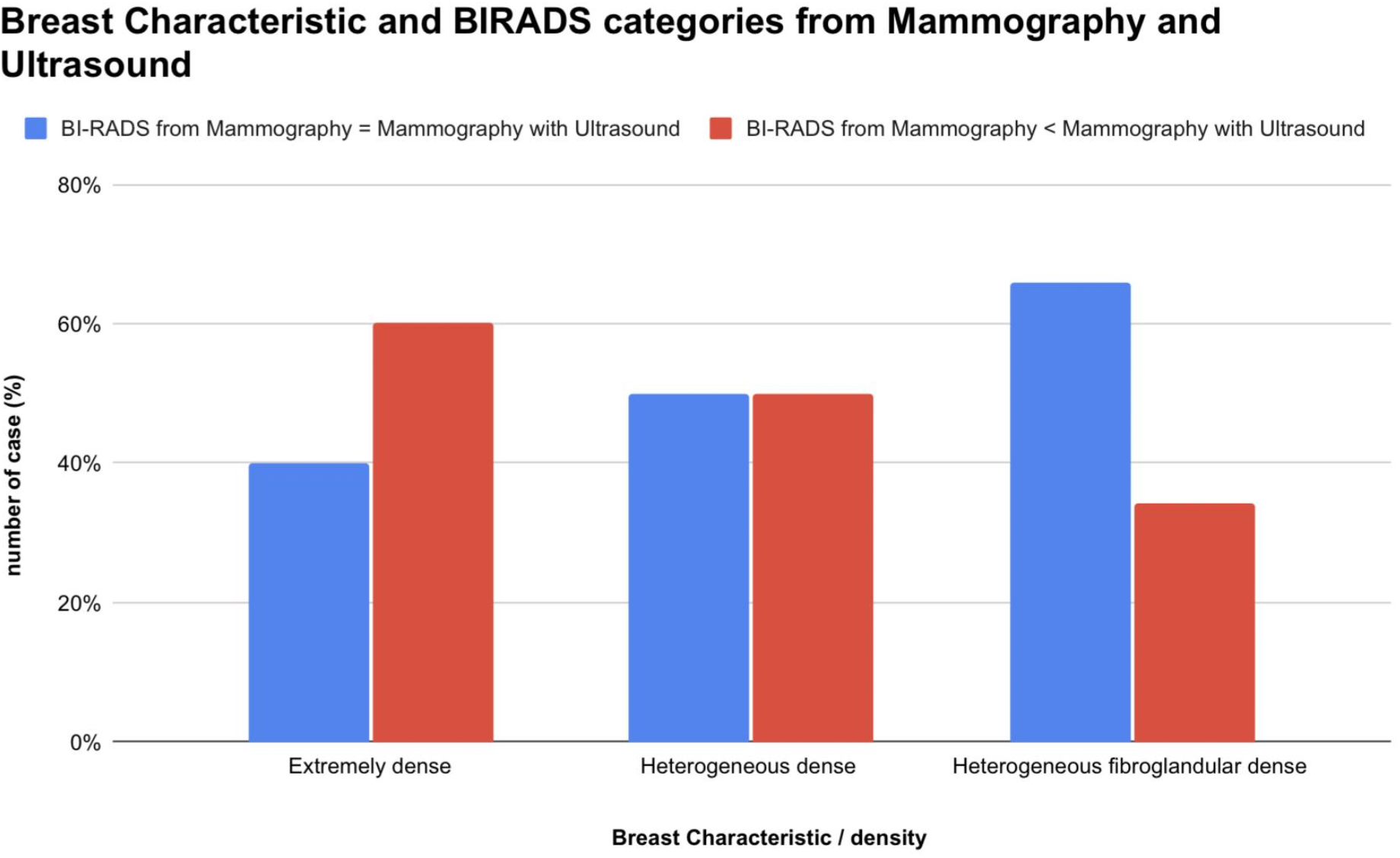

## Discussion

Current clinical practice for breast cancer screening in King Chulalongkorn Memorial Hospital, Bangkok, Thailand are as follows: for women under 40 years old ultrasonography is used with mammography as an adjunct investigation and for women 40 years old and older mammography is used as the main investigation with ultrasonography as an additional investigation. This current practice was adopted from international recommendations without domestic randomized controlled trials to guide actual clinical practice and monitor the effectiveness of the intervention. We question this protocol, as breasts in younger women are correlated with higher breast density, while the inverse relationship holds true with age^16^.

Evidence suggests that dense breasts are strongly associated with increased risk of breast cancer in both premenopausal and postmenopausal women^17^. There should therefore be a corresponding difference to how clinicians approach patients of different breast densities. We suspect that the difference between sensitivity of mammography and ultrasonography stems from differences in breast density rather than age itself. If notable differences are found within different breast densities, it would be advisable for suspected breast cancer patients with dense breasts to suspend the screening examination with mammography and follow up using ultrasonography exclusively, which might improve the cost-effectiveness of nationwide breast cancer screening.

The present study examined the BI-RADS score distribution of middle-aged and elderly Bangkok women, their breast density characteristics, and benefits of adding ultrasound on top of mammogram based on each breast density classification.

As none of our 49 samples presented with BIRAD 4 or 5, we can conclude that there is a very low incidence of suspicious breast abnormalities or malignancy (BI-RADS 4 and 5) detected by mammogram plus ultrasound in randomized middle-aged and older Bangkok women. On the other hand, all of our 49 participants had negative to probably benign findings (BIRAD 1-3). Our result is in accordance with research conducted in the United States, stating that mammography screening for breast cancer yields a detection rate of 3.91 cancer per a thousand examinations^18^. As this our study is based on 49 participants, a larger sample size is needed to confirm the exact incidence of breast cancer in Thai women.

A study concluded that Chinese and Japanese women’s breasts are occupied by dense tissue 20% higher than Caucasian women’s breasts^19^. From our study, we found that the most common breast density in middle-aged and older Thai females is heterogeneous fibroglandular density. Participants with heterogeneous dense breast and extremely dense breast combined accounts for just over 20%. We further split the participants into 2 groups by age: from 30-39 years old and 40-60 years old according to current clinical practice. As we categorized participants in each age group by breast density, we found out that within 7 subjects who are currently less than 40 years old, 6 participants (85.71%) have heterogeneous fibroglandular breast and a single participant (14.29%) with extremely dense breast. For 42 participants within the age group of 40 years old and above, 4 participants (9.52%) have homogeneous dense breasts while the other 38 subjects (90.48%) have heterogeneously dense breasts. Based on this cross-sectional study, we therefore found that younger participants are more likely to present with extremely dense breasts, while presence of heterogeneous dense breasts are higher in older age group. However, a comparison between Thai women’s breast density and breast density of women of other nationalities can’t be made from our current study alone.

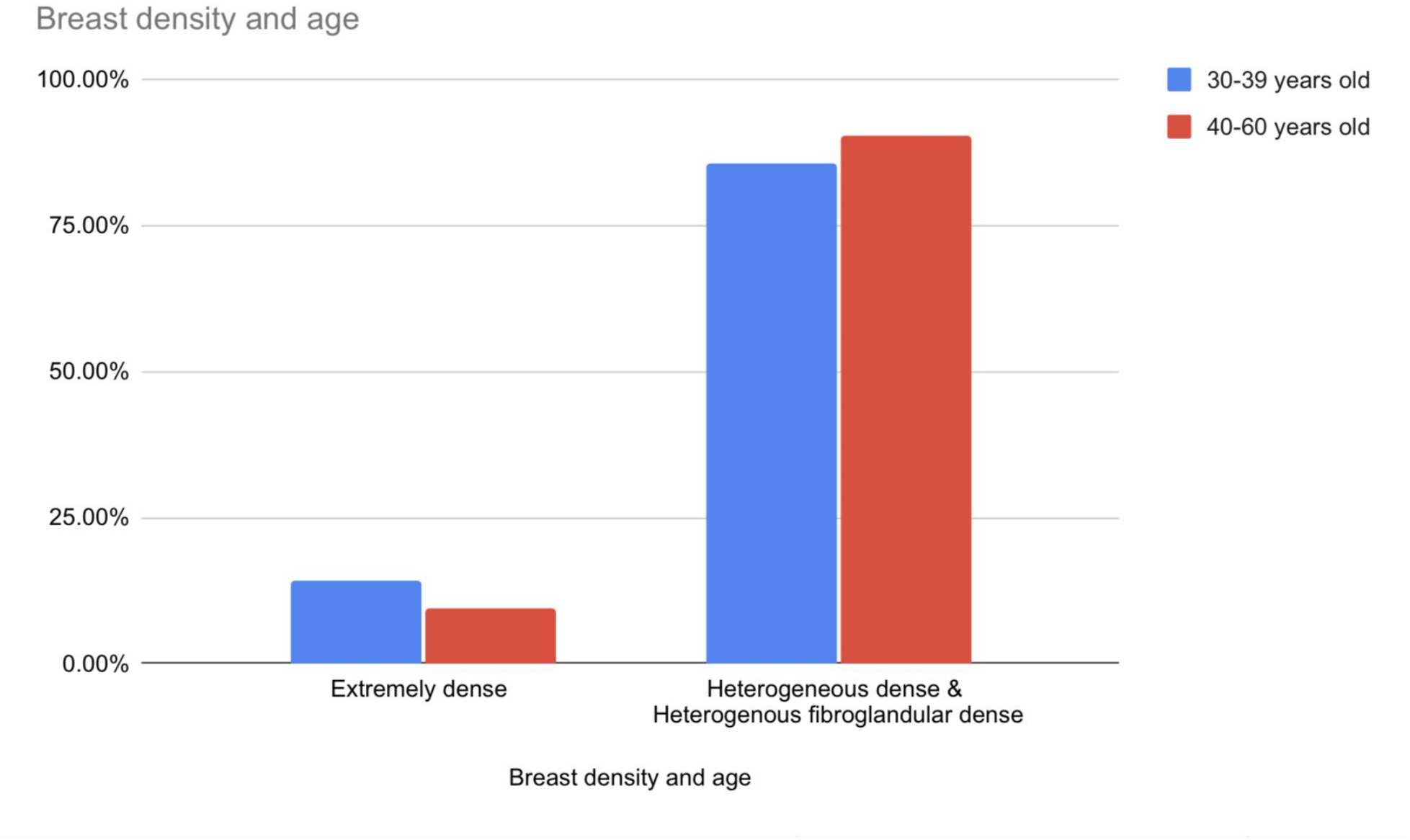

Women with extremely dense breasts present with a 60% BI-RADS score discordance between mammogram alone and mammogram with adjunct ultrasonography. Comparatively, only 50% of subjects with heterogeneous dense breasts and 34.21% of subjects with heterogeneous fibroglandular breasts present such discordance. Our study suggests that women with higher breast density are more likely to be assigned a higher BI-RADS score with adjunct ultrasonography. As such, women with higher breast density are more likely to benefit from the increased sensitivity of higher BI-RADS score which may prompt further necessary investigations or treatments. We recommend differentiating the clinical practice of breast cancer screening between initial screening and the follow-up visits. For the first visit, all women should be screened using both mammography and ultrasonography to ensure the highest detection rate of breast cancer and determine their breast densities. As for follow-up visits, women should be screened according to their breast densities. Women with extremely dense breasts should undergo ultrasonography only and women with heterogeneously dense breasts should be screened with mammography without adjunct ultrasonography. If this practice is implemented, we can hope for increased cost-effectiveness of breast cancer screening in Thailand.

Successful nationwide breast cancer screening comprises 3 main pillars: incidence of breast cancer detected, cost-effectiveness of the screening test and decreased nationwide mortality rate. According to a Thai research published in 2014, breast cancer incidence accounts for 25.6 cases within a population of 100,000^13^. From this figure, screening of breast cancer in asymptomatic Thai general population will yield a very low detection rate. Resulting in the financial need of over 1 million Thai baht for 1 year of quality-adjusted life year (QALY). This represents a very low cost-effectiveness using the current screening procedure. To our knowledge, there is no randomized controlled trial which evaluates the correlation between breast cancer mortality and adjusting breast cancer screening as we proposed. We strongly encourage that a randomized controlled trial be conducted which may change the standard of breast cancer screening practice in Thailand.

## Conclusion

In conclusion, incidence of breast cancer remains very low in middle-aged and older healthy Thai women. The use of adjunct ultrasonography presents a significantly higher diagnostic value, reflected in a higher BI-RADS score detected, in women with higher breast density. We recommend that breast cancer screening practice in Thailand should differentiate between initial visit and follow up visits. For women who screen for breast cancer for the first time, both mammography and ultrasonography should be used to evaluate breast density and provide a higher breast cancer detection rate. For the following visits, we recommend screening in accordance with their breast densities. Women with higher breast densities should undergo ultrasonography only, while women with heterogeneous breast density should be screened by mammography alone. With the use of this recommendation, we can improve cost-effectiveness of breast cancer screening intervention nationally.

## Data Availability

Data produced in the present study are available upon reasonable request to the authors

## Potential conflicts of interests

None

## Notes

### Competing Interest Statement

The authors have declared no competing interest.

### Funding Statement

This study did not receive any funding

### Author Declarations

Data were obtained from the general population who had consented to the mammography and ultrasonographic interventions. Data were collected using an anonymous case report form, without any means of exposing participants personal information. IRB were not deemed necessary by the Chulalongkorn Ethical Review Board.

